# The Impact of Severe Ventricular Remodeling on Quality-of-Life Outcomes after Transcatheter Aortic Valve Replacement

**DOI:** 10.1101/2023.04.28.23289291

**Authors:** Pavan Reddy, Ilan Merdler, Cheng Zhang, Matteo Cellamare, Itsik Ben-Dor, Lowell F. Satler, Toby Rogers, William S. Weintraub, Ron Waksman

## Abstract

**Background:** Among patients with aortic stenosis, severe ventricular remodeling can limit the augmentation of flow with exertion, even after valve intervention. While long-term mortality risk has been shown be elevated in this patient population, the effect on quality of life (QoL) improvement has not been studied. Our objective was to determine the effect of severe ventricular remodeling on QoL outcomes after transcatheter aortic valve replacement (TAVR).

**Methods:** All patients undergoing TAVR from 2011-2021 at our institution were included. Groups were divided according to guideline-recommended cutoffs for left ventricular (LV) wall thickness. Normal or mild wall thickness was designated as non-remodeled (NR) and moderate/severe wall thickness defined the remodeled group (REM). The Kansas City Cardiomyopathy Questionnaire (KCCQ) was utilized to assess QoL and the primary outcome was KCCQ change <5 from baseline to 30 days and 1 year.

**Results:** We analyzed 679 patients (NR: N=389, REM: N=290). Baseline KCCQ overall score [OS] trended higher in the NR group (60.1 vs. 56.6, p=0.053). Echocardiographic baseline differed between NR and REM by septal thickness (1.12 cm vs. 1.44 cm, p<0.001), posterior wall thickness (1.08 cm vs. 1.33 cm, p<0.001), and LV internal diastolic diameter (4.34 cm vs. 4.19 cm, p=0.006). The primary outcome was similar between NR and REM at 30 days (31.6% vs. 28.6%, p=0.449) and at 1 year (27.7% vs. 21.5%, p=0.217). NR and REM experienced similar proportions of worsening or no change, small, moderate, and large improvements in KCCQ score (<5, 5-10, 10-19, ≥20, respectively). Both groups experienced similar domain score changes and NYHA improvement. At one year, NR demonstrated greater reduction in 5MWT time compared to REM (−1.98 s vs. -0.2 s, p=0.028). A subgroup analysis of REM patients did not reveal interaction with cavity size, stroke volume, mitral regurgitation, E/A ratio, valvulo-arterial impedance, or hyperdynamic ejection fraction.

**Conclusions:** Patients with severe ventricular remodeling and aortic stenosis have similar QoL changes after intervention compared to patients without significant remodeling. Findings from this study support aortic valve intervention irrespective of ventricular remodeling.

**What is Known; What the Study Adds:** *What is Known?:*

- Ventricular remodeling occurs due to aortic stenosis, limiting cardiac output even after valve replacement.
- The effect of ventricular remodeling on quality of life after TAVR is unknown.

*What the Study Adds:*

- Quality of life improvement occurred in the majority of patients undergoing TAVR, with and without severe ventricular remodeling, and to a similar degree regardless of remodeling.
- From a quality-of-life perspective, TAVR should remain a viable consideration for patients with severe ventricular remodeling

## Introduction

Among patients with severe aortic stenosis (AS), compensatory left ventricular hypertrophy (LVH), or remodeling, may occur to alleviate high wall stress imposed by outflow obstruction. Maladaptive remodeling leads to small cavity size, fibrosis, and impaired myocardial contractility, causing systolic and diastolic dysfunction, effectively introducing a new and additive disease state.^1^ In severely remodeled ventricles, the ability to augment stroke volume with exertion may be minimal, even after aortic valve replacement, potentially limiting post-procedural symptomatic improvement. Previous studies showed conflicting results regarding the effect of LVH on clinical outcomes after transcatheter aortic valve replacement (TAVR), and few studies assessed the effect on quality of life (QoL),^2-4^ the latter being particularly relevant in elderly patients for whom QoL improvements may be valued more than survival benefits.

QoL assessment using the Kansas City Cardiomyopathy Questionnaire (KCCQ) has been validated for use in the AS population undergoing TAVR.^5^ However, previous predictive models have shown only modest discrimination for poor QoL outcomes and have not reported on the impact of ventricular remodeling.^6^ The objective of this study is, therefore, to determine the effect of severe remodeling on QoL outcomes after TAVR; comprehensive QoL outcomes data will serve to better inform clinicians and patients regarding the benefits and optimal timing of TAVR.

## Methods

All patients who underwent TAVR at our institution between 2011 and 2021 were included in this analysis. Patients were excluded if KCCQ was not performed pre-procedure or at 30-day follow-up visit or if there were insufficient echocardiographic data to determine LVH grade. Cases were also excluded if there was left ventricular (LV) dilation or if ejection fraction (EF) was reduced (EF <52% for men, <54% for women). LVH was graded according to gender-specific cutoffs for septal and posterior wall thickness put forth by the American Society of Echocardiography.^7^ Patients were included in the severely remodeled (REM) cohort if septal or posterior wall thickness met criteria for moderate or severe ventricular hypertrophy; patients with none or mild wall thickness constituted the non-remodeled group (NR).

QoL was assessed using the KCCQ-12. The KCCQ-12 has 12 questions representing 4 domains: symptom frequency, physical limitation, social limitation, and QoL. Compared to the KCCQ-23, the KCCQ-12 has shown good correlation with the aforementioned domain scores and overall score. However, three domain assessments are omitted: symptom burden, symptom stability, and self-efficacy.^8^ The primary outcome of this analysis was a change in KCCQ-overall score (OS) <5 at 30 days and at 1 year. This cutoff value is in accordance with the accepted threshold for minimal clinically important difference in KCCQ-OS.^9^ The KCCQ-OS ranges from 0-100, with higher scores indicating better QoL. Changes in the KCCQ-OS of 5, 10, and 20 points correspond with small, moderate, or large clinical improvements, respectively.^8^ New York Heart Association (NYHA) class and five-meter walk test (5MWT) changes at 30 days and 1 year were also assessed.

This study was approved by the MedStar Washington Hospital Center Institutional Review Board, and all patients provided written informed consent before participation.

### Statistical Analysis

Continuous variables are presented as mean ± standard deviation and categorical variables as proportions. The Student t test and χ^2^ test were used to evaluate the statistical significance between continuous and categorical variables, respectively.

## Results

Between 2011 and 2021, 2038 patients underwent TAVR at our institution, with 1244 having subsequent follow-up. After exclusion for missing data or death, 679 patients were analyzed in the 30-day cohort and 358 patients in the 1-year cohort (Figure 1). Among patients with echocardiography data but who were missing KCCQ assessment, 174 did not have remodeling, and 93 had severe remodeling. Among patients who died, 17 did not have remodeling and 14 had severe remodeling.

**Figure 1.**
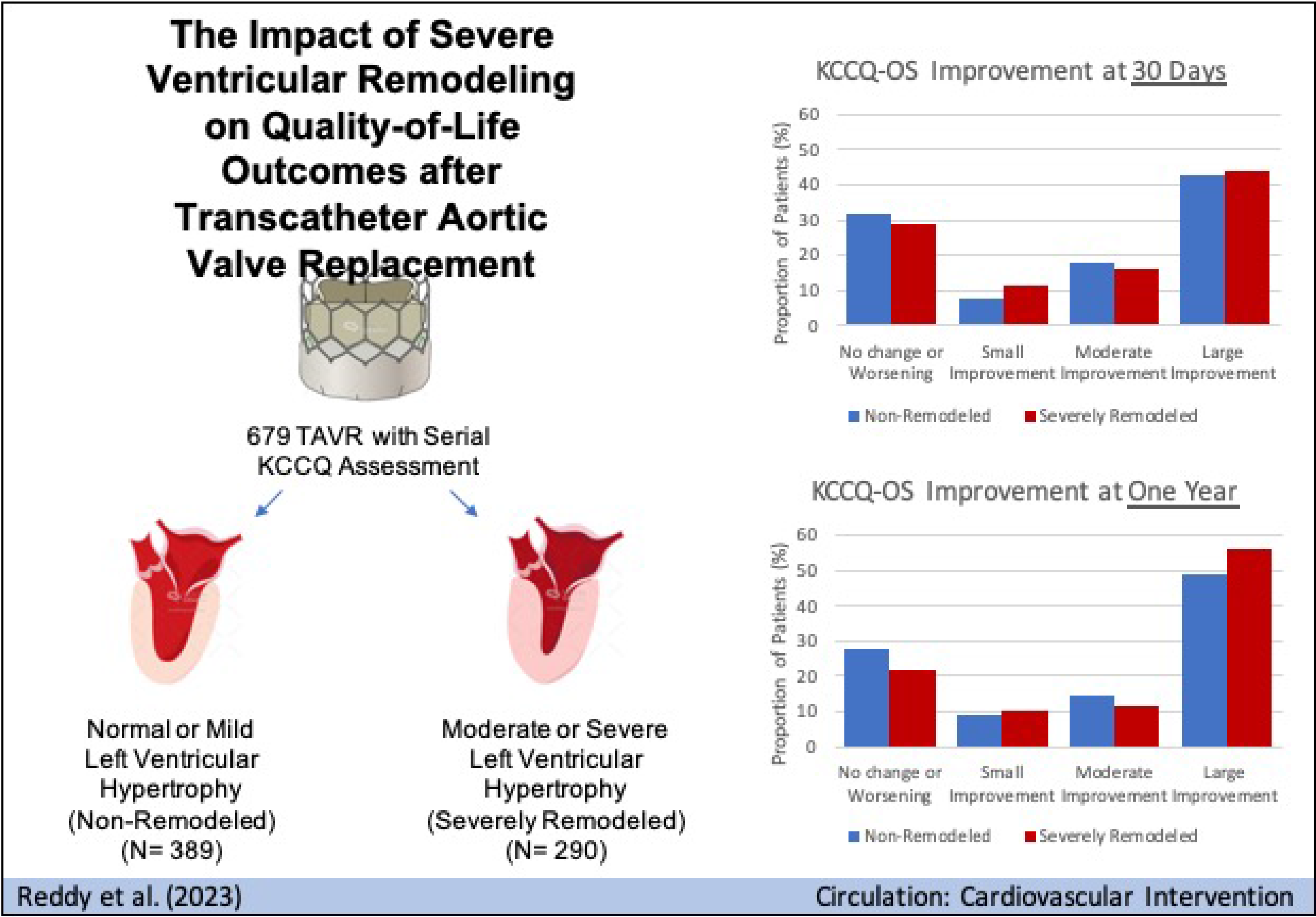
Study Flow Chart F/U: follow-up; KCCQ: Kansas City Cardiomyopathy Questionnaire; LVEF: left ventricular ejection fraction; TAVR: transcatheter aortic valve replacement.

Baseline clinical and echocardiographic characteristics are shown in Table 1 and Table 2, respectively. The mean age of the entire cohort was 79.7 years. NR, compared to REM, patients were more like to be male (54.8% vs. 43.1%) and Caucasian (84.6% vs. 77.9%). Notable clinical differences include a higher prevalence of prior myocardial infarction and permanent pacemaker in the REM group. Pre-procedural symptoms were similar between NR and REM without significant differences in NYHA class, baseline 5MWT, or KCCQ-OS (60.1 vs. 56.6, p=0.053). Baseline echocardiographic measurements show that septal and posterior wall thickness was higher in REM patients, with smaller LV cavity size (4.19 cm vs. 4.34 cm, p=0.006). Pulmonary arterial systolic pressure was also higher in REM patients. EF was statistically higher in REM but with a small, clinically relevant difference (61.6% vs. 60.8%, p=0.036).

**Table 1.** Baseline Clinical Characteristics and Symptoms Assessment.

| **Variable** | **Non-Remodeled**<br>**(N=389)** | **Severe**<br>**Remodeling**<br>**(N=290)** | **P Value** |
| --- | --- | --- | --- |
| Age | 79.8 +/- 8.15 (389) | 79.65 +/- 9.19 (290) | 0.824 |
| Male Gender | 54.8% (213/389) | 43.1% (125/290) | 0.003 |
| White Race | 84.6% (329/389) | 77.9% (226/290) | 0.034 |
| Black Race | 10% (39/389) | 18.6% (54/290) | 0.002 |
| Asian Race | 3.6% (14/389) | 1.4% (4/290) | 0.124 |
| Hispanic Origin | 2.7% (10/370) | 3.5% (10/282) | 0.697 |
| Hypertension | 89.2% (346/388) | 91.7% (266/290) | 0.329 |
| Diabetes | 36.8% (143/389) | 37.5% (108/288) | 0.907 |
| Tobacco Use | 5.9% (23/388) | 8.6% (25/290) | 0.23 |
| Prior Myocardial Infarction | 13.9% (54/389) | 8.7% (25/289) | 0.048 |
| Stroke | 9.5% (37/389) | 12.8% (37/290) | 0.223 |
| Peripheral Arterial Disease | 19.3% (75/389) | 15.9% (46/290) | 0.294 |
| Pacemaker | 6.5% (23/356) | 13.6% (35/257) | 0.004 |
| Coronary Bypass Surgery | 18% (64/356) | 14.7% (38/258) | 0.338 |
| Atrial Fibrillation | 30.3% (118/389) | 34.6% (100/289) | 0.274 |
| Total Albumin | 3.55 +/- 0.51 (387) | 3.55 +/- 0.49 (290) | 0.976 |
| Body Mass Index | 27.95 +/- 5.76 (389) | 30.16 +/- 7.65 (290) | <.001 |
| Body Surface Area (m<sup>2</sup>) | 1.87 +/- 0.25 (389) | 1.91 +/- 0.26 (290) | 0.071 |
| STS Risk Score (%) | 5.09 +/- 4.81 (340) | 5.34 +/- 3.52 (236) | 0.466 |
| **Symptom Baseline** |  |  |  |
| NYHA I | 7.5% (29/388) | 5.9% (17/290) | 0.502 |
| NYHA II | 36.3% (141/388) | 37.6% (109/290) | 0.801 |
| NYHA III | 51.3% (199/388) | 51.4% (149/290) | 1 |
| NYHA IV | 4.9% (19/388) | 5.2% (15/290) | 1 |
| 5 Meter Walk Test (s) | 7.13 +/- 3.41 (316) | 7.56 +/- 3.55 (208) | 0.169 |
| KCCQ12-Overall score | 60.14 +/- 24.4 (389) | 56.55 +/- 23.55 (290) | 0.053 |

Values are % (n/N) or mean ± SD. KCCQ: Kansas City Cardiomyopathy Questionnaire; NYHA: New York Heart Association; STS: Society of Thoracic Surgeons

**Table 2.** Baseline Echocardiographic Characteristics.

| **Variable name** | **Non-Remodeled**<br>**(N=389)** | **Severely**<br>**Remodeled**<br>**(N=290)** | **P Value** |
| --- | --- | --- | --- |
| LV Ejection Fraction (%) | 60.75 ± 4.95<br>(386) | 61.62 ± 5.55<br>(289) | 0.036 |
| Septal Wall Thickness<br>(cm) | 1.12 ± 0.13 (388) | 1.44 ± 0.17<br>(290) | <.001 |
| Posterior Wall Thickness<br>(cm) | 1.08 ± 0.15 (389) | 1.33 ± 0.17<br>(289) | <.001 |
| LVIDd (cm) | 4.34 ± 0.67 (346) | 4.19 ± 0.64<br>(252) | 0.006 |
| LVIDs (cm) | 2.96 ± 0.6 (345) | 2.83 ± 0.62<br>(252) | 0.01 |
| LV Mass (g) | 170.87 ± 50.07<br>(386) | 225.62 ± 60.6<br>(289) | <.001 |
| PASP (mmHg) | 41.65 ± 13.65<br>(257) | 47.15 ± 18.41<br>(193) | 0.001 |
| E/A Ratio | 1.01 ± 0.55 (234) | 1.03 ± 0.55<br>(145) | 0.668 |
| LA volume Index<br>(ml/m<sup>2</sup>) | 37.84 ± 15.82<br>(182) | 40.73 ± 16.83<br>(117) | 0.14 |
| Stroke Volume (mL) | 78.37 ± 19.41<br>(217) | 81.81 ± 26.62<br>(134) | 0.196 |
| Stroke Volume Index<br>(mL/m<sup>2</sup>) | 41.12 ± 10.04<br>(210) | 43.66 ± 16.71<br>(131) | 0.117 |
| Impedance (mm Hg mL-1 m-2) | 4.48 ± 1.09 (190) | 4.5 ± 1.34 (113) | 0.871 |
| Bicuspid Valve | 4.1% (16/389) | 4.1% (12/290) | 1 |
| Aortic Valve Area (cm<sup>2</sup>) | 0.75 ± 0.21 (376) | 0.72 ± 0.16<br>(285) | 0.011 |
| AV Peak Velocity (m/s) | 4.09 ± 0.64 (381) | 4.33 ± 0.65<br>(281) | <.001 |
| AV Peak Gradient<br>(mmHg) | 67.2 ± 21.17<br>(173) | 76.09 ± 23.91<br>(157) | <.001 |
| AV Mean Gradient<br>(mmHg) | 41.58 ± 13.61<br>(382) | 47.39 ± 14.5<br>(288) | <.001 |
| Severe Mitral<br>Regurgitation | 20% (48/240) | 20.5% (36/176) | 1 |
| Severe Aortic<br>Regurgitation | 15.2% (59/389) | 13.8% (40/290) | 0.695 |
| Severe Tricuspid<br>Regurgitation | 11.6% (41/353) | 10.9% (28/257) | 0.883 |

Values are % (n/N) or mean ± SD. AV: aortic valve; LA: left atrial; LV: left ventricular; LVIDd: left ventricular internal diameter diastolic; LVIDs: left ventricular internal diameter systolic; PASP: pulmonary artery systolic pressure.

Similar majorities in NR and REM had clinically significant improvement in KCCQ score after TAVR; at 30 days, the primary endpoint (KCCQ change <5) occurred in 31.6% of NR patients and 28.6% of REM patients (p=0.449), and at 1 year, in 27.7% of NR patients and 21.5% of REM patients (p=0.217) (Table 3). Both groups also experienced similar proportions of no change or worsening, small, moderate, or large improvement in KCCQ (Figure 2). Further breakdown of the individual domain components of KCCQ again show similar changes between groups (Figure 3).

**Table 3.** Quality of Life Outcomes at 30 days and 1 year.

| **Variable** | **Non-Remodeled**<br>**(N=389)** | **Severely**<br>**Remodeled**<br>**(N=290)** | **P**<br>**Value** |
| --- | --- | --- | --- |
| KCCQ-Overall Score at 30d | 77.24 ± 21.4<br>(389) | 73.62 ± 23.14<br>(290) | 0.038 |
| KCCQ-Overall Score at 1y | 80.59 ± 19.47<br>(195) | 78.09 ± 20.59<br>(163) | 0.241 |
| KCCQ Baseline to 30d | 17.1 ± 23.92<br>(389) | 17.07 ± 25.65<br>(290) | 0.991 |
| KCCQ Baseline to 1y | 21.16 ± 25.94<br>(195) | 22.47 ± 23.33<br>(163) | 0.613 |
| KCCQ Increase < 5 at 30d | 31.6% (123/389) | 28.6% (83/290) | 0.449 |
| KCCQ Increase < 5 at 1y | 27.7% (54/195) | 21.5% (35/163) | 0.217 |
| NYHA I at 30d | 44.1% (162/367) | 36.3% (98/270) | 0.056 |
| NYHA II at 30d | 42% (154/367) | 44.1% (119/270) | 0.652 |
| NYHA III at 30d | 12.8% (47/367) | 18.5% (50/270) | 0.061 |
| NYHA IV at 30d | 1.1% (4/367) | 1.1% (3/270) | 1 |
| NYHA I at 1y | 34.2% (51/149) | 30.2% (39/129) | 0.561 |
| NYHA II at 1y | 32.9% (49/149) | 37.2% (48/129) | 0.53 |
| NYHA III at 1y | 29.5% (44/149) | 29.5% (38/129) | 1 |
| NYHA IV at 1y | 3.4% (5/149) | 3.1% (4/129) | 1 |
| 5MWT Baseline to 30d (s) | -0.63 ± 3.15<br>(179) | -0.5 ± 2.41<br>(105) | 0.695 |
| 5MWT Baseline to 1 y (s) | -1.98 ± 4.04<br>(64) | -0.2 ± 4.8 (60) | 0.028 |

Values are % (n/N) or mean ± SD. KCCQ: Kansas City Cardiomyopathy Questionnaire; NYHA: New York Heart Association; 5MWT: 5-meter walk test.

**Figure 2.**
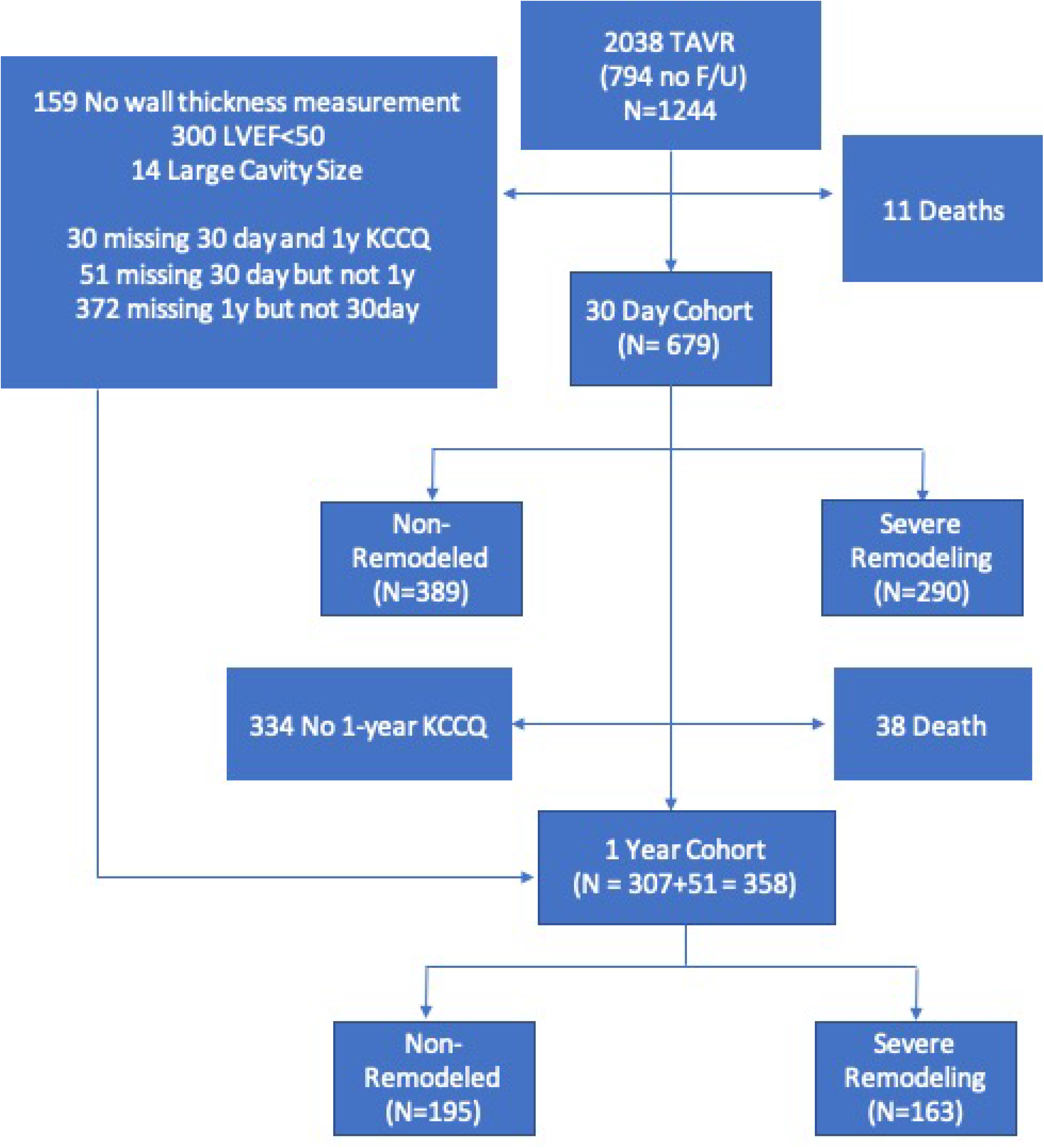
Analysis of Clinically Meaningful Changes in Kansas City Cardiomyopathy Questionnaire. A. Improvement in KCCQ at 30 days. B. Improvement in KCCQ at 1 year. KCCQ-OS: Kansas City Cardiomyopathy Questionnaire overall score.

**Figure 3.**
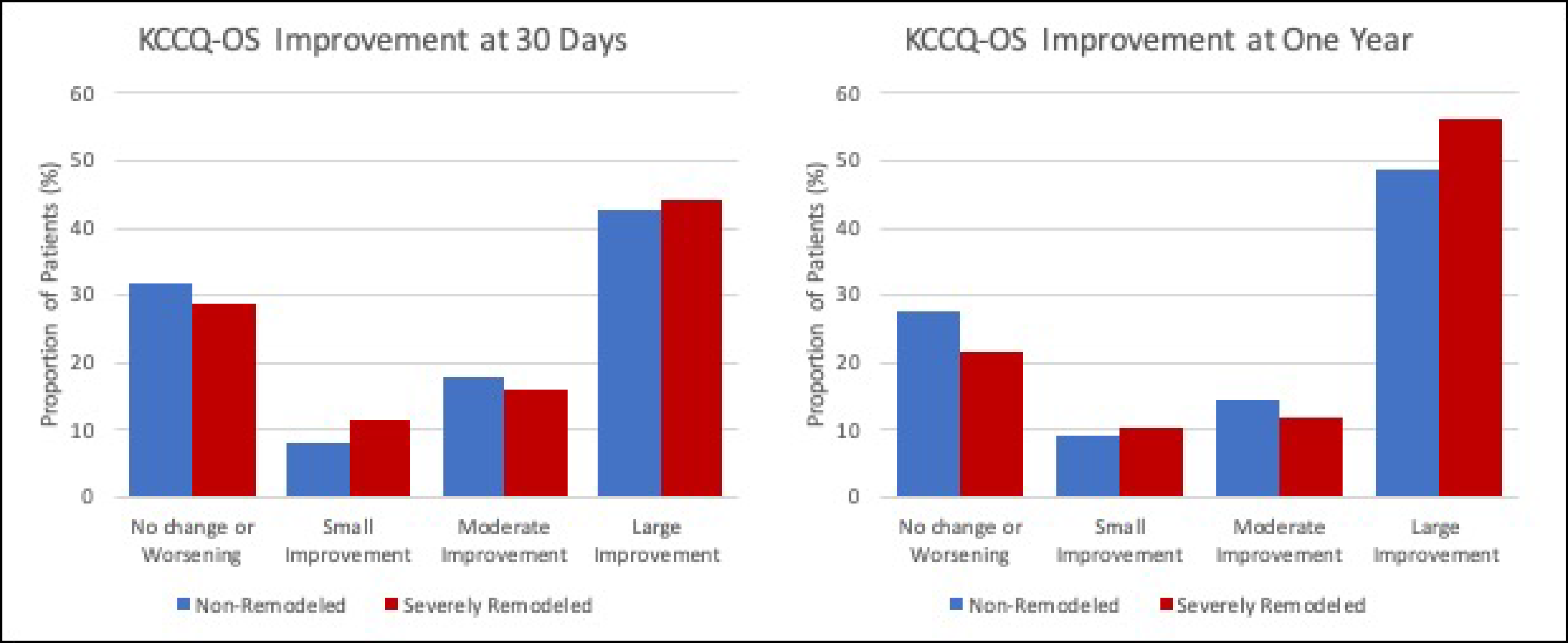
Kansas City Cardiomyopathy Questionnaire Domain Analysis. A. KCCQ Overall Score at baseline, 30 days and 1 year. B. KCCQ Physical Limitation Score at baseline, 30 days and 1 year. C. KCCQ Symptom Frequency Score at baseline, 30 days and 1 year. D. KCCQ Quality of Life Score at baseline, 30 days and 1 year. E. KCCQ Social Limitation Score at baseline, 30 days and 1 year. KCCQ: Kansas City Cardiomyopathy Questionnaire.

The NR and REM groups demonstrated similar proportions of patients at NYHA class I-IV at 30 days and 1 year (Table 3). The degree of NYHA class improvement was also similar between groups (Figure 4). While 5MWT changes were similar at 30 days, at one year, NR demonstrated greater reduction in 5MWT time compared to REM (−1.98 s vs. -0.2 s, p=0.028). A subgroup analysis of patients with REM did not reveal interaction with cavity size, stroke volume, mitral regurgitation, E/A ratio, valvulo-arterial impedance or hyperdynamic ejection fraction (Table 4).

**Table 4.** Subgroup Analysis of KCCQ Improvement Among Patients with Severe Remodeling.

| **Variable** | **Normal Cavity** | **Small Cavity** | **P Value** |
| --- | --- | --- | --- |
| KCCQ Baseline | 57.2 +/- 24.65<br>(147) | 55.87 +/- 22.42<br>(143) | 0.631 |
| KCCQ delta 30 days | 18.21 +/- 27.24<br>(147) | 15.9 +/- 23.93<br>(143) | 0.444 |
| KCCQ delta 1 year | 22.02 +/- 21.67<br>(72) | 22.84 +/- 24.68<br>(91) | 0.822 |
|  | **Normal SV Index** | **Low SV Index** |  |
| KCCQ Baseline | 55.98 +/- 22.32<br>(98) | 57.4 +/- 29.2 (33) | 0.8 |
| KCCQ delta 30 days | 16.4 +/- 29.03<br>(98) | 15.99 +/- 23.13<br>(33) | 0.934 |
| KCCQ delta 1 year | 23.46 +/- 23 (32) | 18.78 +/- 22.32<br>(11) | 0.559 |
|  | **Non-Severe MR** | **Severe MR** |  |
| KCCQ Baseline | 54.6 +/- 22.68<br>(140) | 52.97 +/- 23.02<br>(36) | 0.705 |
| KCCQ delta 30 days | 17.27 +/- 24.44<br>(140) | 19.05 +/- 33.43<br>(36) | 0.766 |
| KCCQ delta 1 year | 22.89 +/- 23.92<br>(86) | 24.84 +/- 25.3<br>(24) | 0.737 |
|  | **E/A < 1** | **E/A ≥ 1** |  |
| KCCQ Baseline | 53.04 +/- 24.92<br>(50) | 60.76 +/- 23.21<br>(95) | 0.073 |
| KCCQ delta 30 days | 17.78 +/- 25.27<br>(50) | 16.29 +/- 25.46<br>(95) | 0.136 |
| KCCQ delta 1 year | 22.94 +/- 23.99<br>(24) | 22.89 +/- 22.36<br>(46) | 0.993 |
|  | **Normal Impedance** | **High Impedance** |  |
| KCCQ Baseline | 56.1 +/- 26.88<br>(29) | 56.48 +/- 23.27<br>(84) | 0.946 |
| KCCQ delta 30 days | 17.5 +/- 28.06<br>(29) | 14.26 +/- 28.62<br>(84) | 0.596 |
| KCCQ delta 1 year | 15.8 +/- 20.32 (3) | 22.82 +/- 22.99<br>(32) | 0.618 |
|  | **Normal EF** | **Hyperdynamic EF** |  |
| KCCQ Baseline | 56.17 +/- 23.51<br>(250) | 58.02 +/- 23.54<br>(39) | 0.649 |
| KCCQ delta 30 days | 18.02 +/- 25.45<br>(250) | 12.11 +/- 26.07<br>(39) | 0.193 |
| KCCQ delta 1 year | 22.29 +/- 22.99<br>(131) | 23.22 +/- 25.03<br>(32) | 0.85 |

Values are mean ± SD. Small cavity is defined by left ventricular internal diastolic diameter < 4.2cm in men and 3.8cm in women. Low SV index is defined as <35 ml/m<sup>2</sup>. High impedance is defined by > 5 mm Hg mL<sup>-1</sup> m<sup>-2</sup>. Hyperdynamic EF is defined as >70%. EF: ejection fraction; KCCQ: Kansas City Cardiomyopathy Questionnaire; MR: mitral regurgitation; SV: stroke volume.

**Figure 4.**
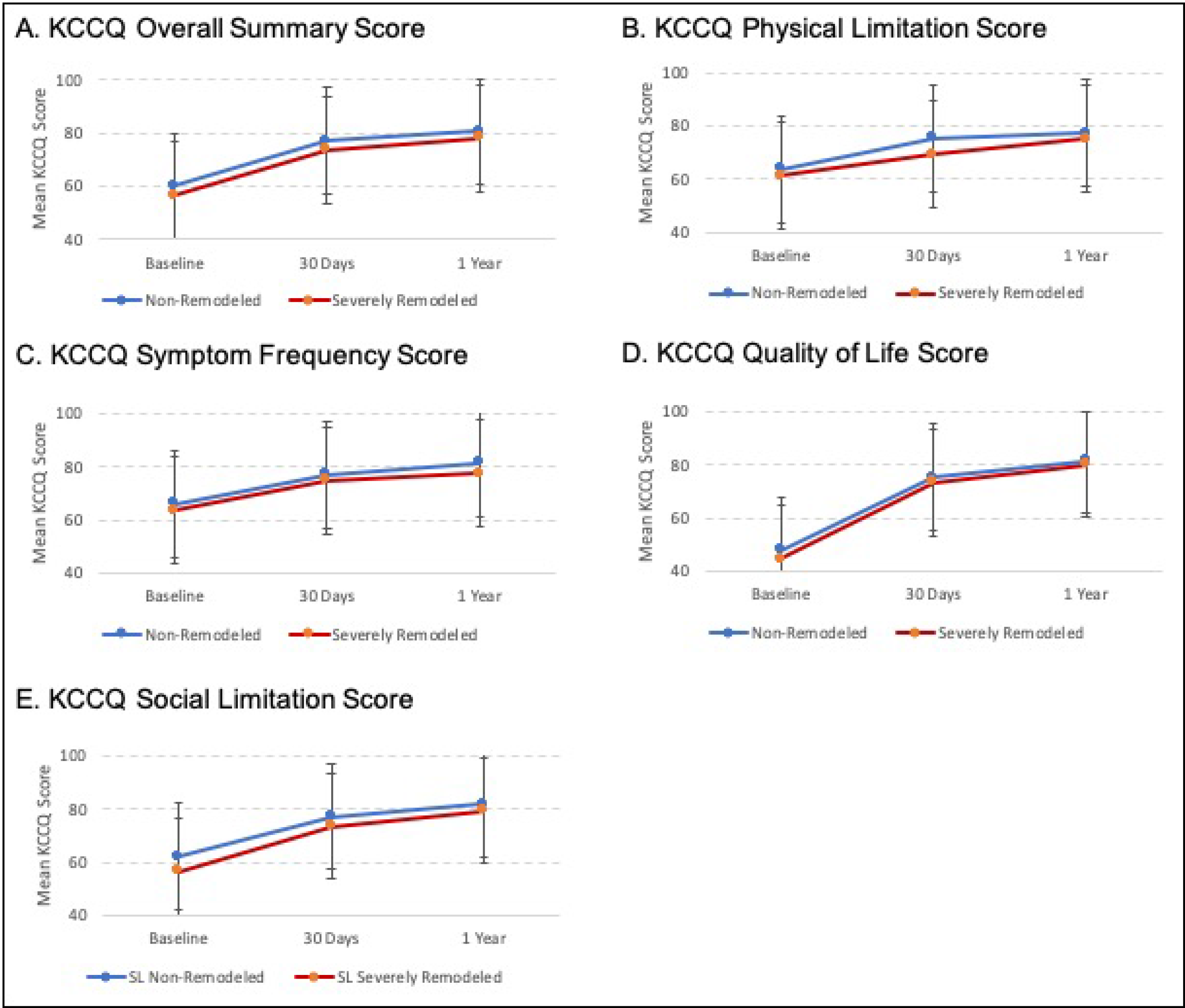
New York Heart Association Class Improvement. A. NYHA Improvement at 30 days. B. NYHA Improvement at 1 year. NYHA: New York Heart Association.

## Discussion

There are several salient findings from this study: 1) Patients with AS and severe ventricular remodeling have similar improvement in QoL after TAVR, as assessed by KCCQ, at 30 days and at 1 year compared to patients without severe remodeling; 2) changes in KCCQ domain scores were not shown to differ significantly between groups; and 3) KCCQ changes in both cohorts were consistent with changes in NYHA class.

With innovative medical therapies now permitting intervention in elderly and highly morbid patients, QoL assessment has become increasingly important.^10,11^ Patients with AS have been shown to experience improved quality of life after AVR as the ability to augment cardiac output with increasing metabolic demands is restored. Relieved of fixed severe obstruction, the ventricle can empty more completely, increasing stroke volume.^12^ However, among patients with severely remodeled ventricles, it is plausible that TAVR, in this late stage, does not offer the same opportunity to augment flow given that the stiff and small-cavitied ventricle cannot accommodate more volume; cardiac output increases only slightly by tachycardia.^12^ The findings of our study to some extent contradict this physiologic assumption; patients with severely remodeled ventricles continued to have significant, sometimes very large, improvements in overall KCCQ and across all domains. Importantly, prior studies have shown that subjective improvement in QoL translates to reduction in mortality and hospitalization rates.^13^ Given the elevated long-term mortality risk and lack of randomized trial data to confirm the benefit of intervention in late-stage AS, QoL results from our study continue to support TAVR for patients with severe remodeling.^3,14^

Findings from our study suggest that significant ventricular remodeling should not preclude TAVR. However, there remains uncertainty regarding which patient attributes are relevant in predicting symptom improvement after intervention. Of note, rarely do singular clinical or hemodynamic metrics accurately predict the presence or degree of AS-related symptoms, prominently exemplified by inconsistent symptomology related to aortic valve gradient. Notably, KCCQ changes in response to TAVR have been persistently difficult to predict despite extensive modeling;^13,15^ this may be due in part to the variability inherent in subjective answers or could otherwise indicate difficulty in capturing more sensitive variables to assess and predict improvement in QoL. Measures that combine structural and hemodynamic characteristics to represent the global impact of AS may prove more useful in predicting QoL outcomes compared to traditional risk factors. Cardiac magnetic resonance (CMR) is apt in this respect, as it provides accurate structural measurements while also revealing tissue properties and even hemodynamic patterns.^16,17^ Tissue fibrosis, of note, has been correlated with both symptoms and prognosis in AS.^1^ Valvulo-arterial impendence is another measure that may gauge the whole syndrome of AS by intentionally accounting for the interaction among ventricle, valve, and vasculature.^18^

Several limitations should be considered when drawing conclusions from this study: 1) Determination of remodeling relied on linear measurement of wall thickness rather than a more comprehensive evaluation, such as that offered by CMR; this may cause non-differential misclassification of cases, biasing the analysis toward the reported null result. 2) We present one-year QoL assessment, but long-term changes in QoL were not available; given that reverse remodeling may occur after AVR, QoL improvements may be potentially understated.^19^ 3) The subgroup analysis of KCCQ changes among patients with severe remodeling is limited by sample size and is, therefore, exploratory in nature.

In conclusion, patients with severe ventricular remodeling and AS have similar QoL changes after intervention compared to patients without severe remodeling. Findings from this study support continued aortic valve intervention irrespective of ventricular remodeling.

## Data Availability

The data that support the findings of this study are available on reasonable request to the corresponding author.

## Abbreviations and Nonstandard Acronyms

AS: aortic stenosis
EF: ejection fraction
KCCQ: Kansas City Cardiomyopathy Questionnair
LV: left ventricular
LVH: left ventricular hypertrophy
NR: non-remodeled group
NYHA: New York Heart Association
QoL: quality of life
REM: remodeled
TAVR: transcatheter aortic valve replacement

---

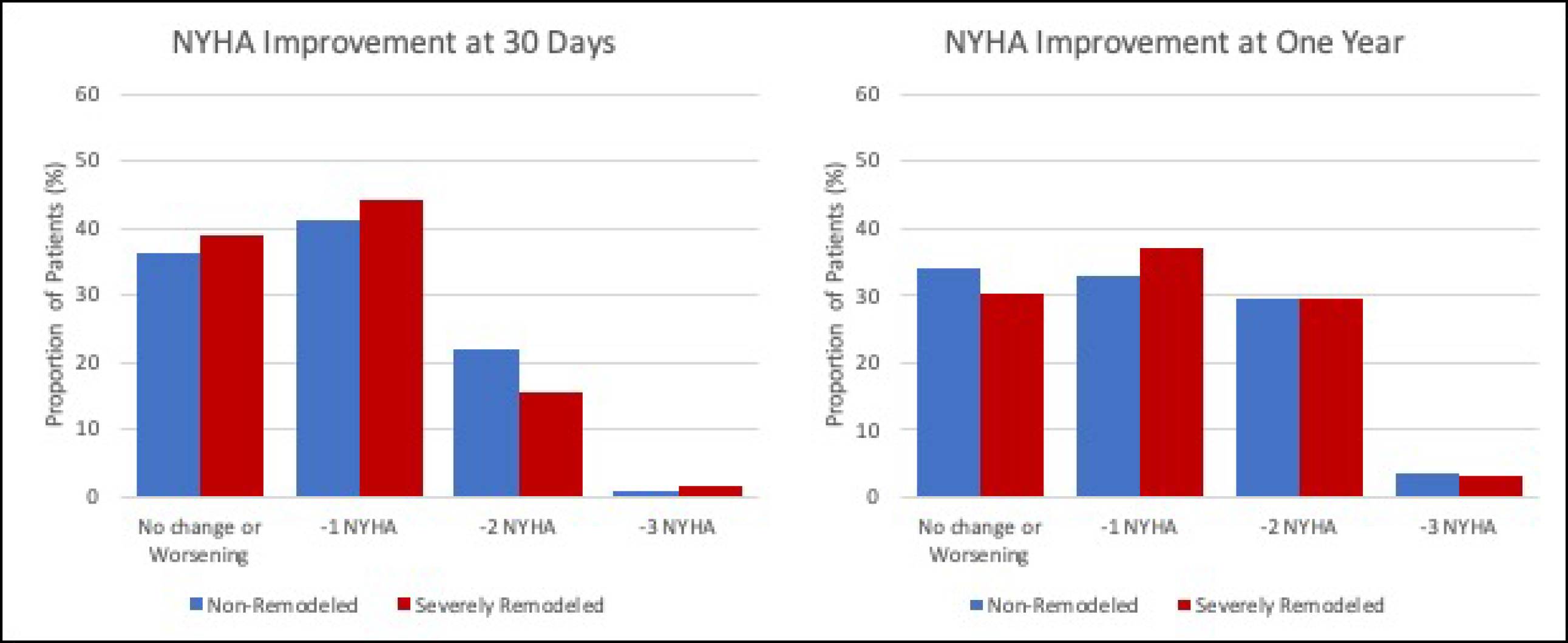

## References

1. Weidemann F, Herrmann S, Störk S, Niemann M, Frantz S, Lange V, Beer M, Gattenlöhner S, Voelker W, Ertl G, et al. Impact of myocardial fibrosis in patients with symptomatic severe aortic stenosis. Circulation. 2009;120:577–584. doi:10.1161/circulationaha.108.847772

2. Varshney AS, Manandhar P, Vemulapalli S, Kirtane AJ, Mathew V, Shah B, Lowenstern A, Kosinski AS, Kaneko T, Thourani VH, et al. Left Ventricular Hypertrophy Does Not Affect 1-Year Clinical Outcomes in Patients Undergoing Transcatheter Aortic Valve Replacement. JACC Cardiovasc Interv. 2019;12:373–382. doi:10.1016/j.jcin.2018.11.013

3. Gonzales H, Douglas PS, Pibarot P, Hahn RT, Khalique OK, Jaber WA, Cremer P, Weissman NJ, Asch FM, Zhang Y, et al. Left Ventricular Hypertrophy and Clinical Outcomes Over 5 Years After TAVR: An Analysis of the PARTNER Trials and Registries. JACC Cardiovasc Interv. 2020;13:1329–1339. doi:10.1016/j.jcin.2020.03.011

4. Dahiya G, Kyvernitakis A, Joshi AA, Lasorda DM, Bailey SH, Raina A, Biederman RWW, Kanwar MK. Impact of transcatheter aortic valve replacement on left ventricular hypertrophy, diastolic dysfunction and quality of life in patients with preserved left ventricular function. Int J Cardiovasc Imaging. 2021;37:485–492. doi:10.1007/s10554-020-02015-z

5. Arnold SV, Spertus JA, Lei Y, Allen KB, Chhatriwalla AK, Leon MB, Smith CR, Reynolds MR, Webb JG, Svensson LG, et al. Use of the Kansas City Cardiomyopathy Questionnaire for monitoring health status in patients with aortic stenosis. Circ Heart Fail. 2013;6:61–67. doi:10.1161/circheartfailure.112.970053

6. Arnold SV, Afilalo J, Spertus JA, Tang Y, Baron SJ, Jones PG, Reardon MJ, Yakubov SJ, Adams DH, Cohen DJ. Prediction of Poor Outcome After Transcatheter Aortic Valve Replacement. J Am Coll Cardiol. 2016;68:1868–1877. doi:10.1016/j.jacc.2016.07.762

7. Lang RM, Badano LP, Mor-Avi V, Afilalo J, Armstrong A, Ernande L, Flachskampf FA, Foster E, Goldstein SA, Kuznetsova T, et al. Recommendations for cardiac chamber quantification by echocardiography in adults: an update from the American Society of Echocardiography and the European Association of Cardiovascular Imaging. J Am Soc Echocardiogr. 2015;28:1–39.e14. doi:10.1016/j.echo.2014.10.003

8. Spertus JA, Jones PG, Sandhu AT, Arnold SV. Interpreting the Kansas City Cardiomyopathy Questionnaire in Clinical Trials and Clinical Care: JACC State-of-the-Art Review. J Am Coll Cardiol. 2020;76:2379–2390. doi:10.1016/j.jacc.2020.09.542

9. Spertus J, Peterson E, Conard MW, Heidenreich PA, Krumholz HM, Jones P, McCullough PA, Pina I, Tooley J, Weintraub WS, et al. Monitoring clinical changes in patients with heart failure: a comparison of methods. Am Heart J. 2005;150:707–715. doi:10.1016/j.ahj.2004.12.010

10. U.S. Food and Drug Administration. Treatment for Heart Failure: Endpoints for Drug Development Guidance for Industry. Bethesda, MD: Food and Drug Administration. June 2019. https://www.fda.gov/regulatory-information/search-fda-guidance-documents/treatment-heart-failure-endpoints-drug-development-guidance-industry (Accessed January 10, 2023).

11. U.S. Food and Drug Administration. Statement from FDA Commissioner Scott Gottlieb, M.D., on new steps to advance medical device innovation and help patients gain faster access to beneficial technologies. [News release]. October 24, 2017. https://www.fda.gov/news-events/press-announcements/statement-fda-commissioner-scott-gottlieb-md-new-steps-advance-medical-device-innovation-and-help (Accessed January 10, 2023).

12. Carabello B. Chapter 5: Left Ventricular Adaptation to Pressure and/or Volume Overload. In: Otto CM, Bonow RO, eds. Valvular Heart Disease: A Companion to Braunwald’s Heart Disease. 3rd ed. Saunders/Elsevier; 2009:55–61. doi:10.1016/B978-1-4160-5892-2.00004-0

13. Arnold SV, Spertus JA, Vemulapalli S, Dai D, O’Brien SM, Baron SJ, Kirtane AJ, Mack MJ, Green P, Reynolds MR, et al. Association of Patient-Reported Health Status With Long-Term Mortality After Transcatheter Aortic Valve Replacement: Report From the STS/ACC TVT Registry. Circ Cardiovasc Interv. 2015;8:e002875. doi:10.1161/circinterventions.115.002875

14. Saito T, Inohara T, Yoshijima N, Yashima F, Tsuruta H, Shimizu H, Fukuda K, Naganuma T, Mizutani K, Yamawaki M, et al. Small Left Ventricle and Clinical Outcomes After Transcatheter Aortic Valve Replacement. J Am Heart Assoc. 2021;10:e019543. doi:10.1161/jaha.120.019543

15. Arnold SV, Cohen DJ, Dai D, Jones PG, Li F, Thomas L, Baron SJ, Frankel NZ, Strong S, Matsouaka RA, et al. Predicting Quality of Life at 1 Year After Transcatheter Aortic Valve Replacement in a Real-World Population. Circ Cardiovasc Qual Outcomes. 2018;11:e004693. doi:10.1161/circoutcomes.118.004693

16. Komoriyama H, Kamiya K, Nagai T, Oyama-Manabe N, Tsuneta S, Kobayashi Y, Kato Y, Sarashina M, Omote K, Konishi T, et al. Blood flow dynamics with four-dimensional flow cardiovascular magnetic resonance in patients with aortic stenosis before and after transcatheter aortic valve replacement. J Cardiovasc Magn Reson. 2021;23:81. doi:10.1186/s12968-021-00771-y

17. Kwak S, Everett RJ, Treibel TA, Yang S, Hwang D, Ko T, Williams MC, Bing R, Singh T, Joshi S, et al. Markers of Myocardial Damage Predict Mortality in Patients With Aortic Stenosis. J Am Coll Cardiol. 2021;78:545–558. doi:10.1016/j.jacc.2021.05.047

18. Nuis RJ, Goudzwaard JA, de Ronde-Tillmans M, Kroon H, Ooms JF, van Wiechen MP, Geleijnse ML, Zijlstra F, Daemen J, Van Mieghem NM, et al. Impact of Valvulo-Arterial Impedance on Long-Term Quality of Life and Exercise Performance After Transcatheter Aortic Valve Replacement. Circ Cardiovasc Interv. 2020;13:e008372. doi:10.1161/circinterventions.119.008372

19. Une D, Mesana L, Chan V, Maklin M, Chan R, Masters RG, Mesana TG, Ruel M. Clinical Impact of Changes in Left Ventricular Function After Aortic Valve Replacement: Analysis From 3112 Patients. Circulation. 2015;132:741–747. doi:10.1161/circulationaha.115.015371

